# Acceptability and feasibility of strategies to shield the vulnerable during the COVID-19 outbreak: a qualitative study in six Sudanese communities

**DOI:** 10.1101/2020.12.14.20248160

**Authors:** Nada Abdelmagid, Salma A.E. Ahmed, Nazik Nurelhuda, Israa Zainalabdeen, Aljaile Ahmed, Mahmoud Ali Fadlallah, Maysoon Dahab

**Affiliations:** London School of Hygiene and Tropical Medicine, (Department of Infectious Disease Epidemiology), London, (London), United Kingdom; Independent public health researcher, Khartoum, (Khartoum), Sudan; University of Khartoum, Faculty of Dentistry, Khartoum, (Khartoum), Sudan; Y-PEER Sudan, Khartoum, (Khartoum), Sudan; Asian Institute of Technology, Bangkok, (Bangkok), Thailand; Public Health Institute (PHI), Khartoum, (Khartoum), Sudan; Sudan COVID-19 Research Group

**Keywords:** Shielding, COVID-19, Sudan, High-risk, Resource-poor, Acceptability, Feasibility, Participatory analysis

## Abstract

**Background:** Shielding of high-risk groups from coronavirus disease (COVID-19), either within their households or safe communal structures, has been suggested as a realistic alternative to severe movement restrictions in response to the COVID-19 epidemic in low-income countries. To our knowledge, this concept has not been tested or evaluated in resource-poor settings. This study aimed to explore the acceptability and feasibility of strategies to shield persons at higher risk of severe COVID-19 outcomes, during the COVID-19 epidemic in six communities in Sudan.

**Methods:** We purposively sampled participants from six communities, illustrative of urban, rural and forcibly-displaced settings. In-depth telephone interviews were held with 59 members of households with one or more members at higher risk of severe COVID-19 outcomes. Follow-up interviews were held with 30 community members after movement restrictions were eased across the country. All interviews were audio-recorded, transcribed verbatim, and analysed using a two-stage deductive and inductive thematic analysis.

**Results:** Most participants were aware that some people are at higher risk of severe COVID-19 outcomes but were unaware of the concept of shielding. Most participants found shielding acceptable and consistent with cultural inclinations to respect elders and protect the vulnerable. However, extra-household shielding arrangements were mostly seen as socially unacceptable. Participants reported feasibility concerns related to the social isolation of shielded persons and loss of income for shielding families. The acceptability and feasibility of shielding strategies were reduced after movement restrictions were eased, as participants reported lower perception of risk in their communities and increased pressure to comply with social commitments outside the house.

**Conclusion:** Shielding is generally acceptable in the study communities. Acceptability is influenced by feasibility, and by contextual changes in the epidemic and associated policy response. The promotion of shielding should capitalise on the cultural and moral sense of duty towards elders and vulnerable groups. Communities and households should be provided with practical guidance to implement feasible shielding options. Households must be socially, psychologically and financially supported to adopt and sustain shielding effectively.

## Background

On January 30, 2020, the World Health Organization (WHO) declared the coronavirus disease (COVID-19) outbreak a “Public Health Emergency of International Concern” [1]. The first COVID-19 case in Sudan was reported on March 13, 2020. The Government of Sudan immediately declared a national emergency [2] and enforced schools and universities closures [3]. Bans of mass gatherings [4] and border closures [5] followed shortly after. However, a reprieve in late March to let in stranded Sudanese travellers [6] may have resulted in the importation of more COVID-19 cases [7]. Following increasingly restrictive dusk-to-dawn curfews in Khartoum, the capital, a lockdown was started on April 18 2020 [8], and, soon after, movement restrictions were extended to other parts of the country [7].

From the beginning, Sudan adopted a test, track and quarantine strategy for containment of the epidemic [9]. Like other African countries, Sudan acted early, instituting a lockdown when the country had recorded 66 confirmed COVID-19 cases and 10 deaths [10].

COVID-19 comes to Sudan at a time of heightened political and economic fragility, after emerging from decades of authoritarian rule. The response in Sudan has been challenged by a pre-pandemic fragile health system, weak social protection systems and difficulties in accessing global emergency funding for COVID-19 [11]. Likely, the true magnitude of the COVID-19 epidemic in Sudan is significantly underestimated by official statistics, as testing capacity is limited and fraught with operational and logistical difficulties [12]. Moreover, among 28 African countries, Sudan has the lowest levels of adherence to crucial prevention measures, such as mask-wearing, physical distancing and compliance with movement restrictions [13], which may partially be due to severe economic hardship faced by the population during the epidemic.

In low-income countries like Sudan, prevention is key to mitigating the impact of COVID-19. Shielding of high-risk individuals, either within their homes or in safe communal structures, has been suggested as an alternative to widely-enforced social distancing measures such as lockdowns. The approach recognises that, in these settings, high-risk individuals, especially the elderly, usually co-reside with young people and children in large households [14]. While shielding does not significantly affect transmission dynamics [15], focusing limited resources on shielding higher-risk individuals aims to reduce pressure on health systems, prevent COVID-19 deaths and allow economic activities to resume. Through mathematical modelling, shielding seems to be effective at reducing mortality from COVID-19, particularly when coupled with moderate social distancing and high levels of self-isolation of those with mild COVID-19 symptoms [15].

The shielding approach is premised on sound epidemiological principles that aim to reduce the number and duration of effective contacts among high-risk groups and therefore, their risk of infection [16]. However, the practical application of shielding in resource-poor settings may be challenging, and effective shielding strategies will need to be developed and adapted locally, in a manner that is acceptable and feasible to the different contexts [17-19]. In this study, we explored the acceptability and feasibility of shielding strategies in six illustrative communities in Sudan, to provide information to government and non-government COVID-19 responders in Sudan seeking to protect high-risk groups during the epidemic.

## Methods

### Study design and setting

This qualitative study was conducted in participation with Sudan’s Youth Peer Education Network (Y-PEER Sudan), a nationwide network of trained youth volunteers active in promoting health and youth participation. At the time of the study, Y-PEER Sudan was conducting awareness-raising campaigns on COVID-19 prevention in communities across Sudan. We selected six communities in five Sudanese states, illustrative of different contextual settings. In Khartoum state, we selected Umbadda district, a high-density poor urban population, and Tuti Island, a close-knit agricultural community in the heart of the capital. We also selected two other urban communities, El-Obeid, a high-density city in North Kordofan State, and Damazin North, a peri-urban setting on the shared country border with South Sudan. Finally, we selected one rural village in the south of Gezira state, and one internally displaced person’s (IDP) camp in South Darfur (Dereij IDP camp). At the time of planning in early April 2020, only Khartoum state had reported confirmed COVID-19 cases and deaths.

The study was conducted remotely, complying with safety precautions and movement restrictions due to the COVID-19 epidemic. Youth peers were trained remotely, through pre-recorded video and audio training sessions on qualitative research, shielding strategies, informed consent and study procedures, developed and delivered by the first author (NA). Training materials were shared via social media platforms, and this was followed by a live question and answer social media session with three co-authors (NA, NN and MD).

### Study participants

We interviewed two groups of participants in the six study sites at different times: once during periods of movement restriction enforced in April 2020 and once after restrictions were eased in July 2020. To be eligible, participants in the first round of interviews had to be an adult member of a household that had at least one member at higher risk of severe COVID-19 outcomes. Participants in the second round of interviews were any adults in the community. For this study, we defined those at high risk as persons aged 60 years or more, or persons of any age with one of the following chronic diseases: hypertension, diabetes, cardiovascular disease, chronic respiratory disease, chronic kidney failure, cancer and persons with immunosuppressive diseases or receiving immunosuppressant therapy.

Youth peers purposively identified eligible participants through their existing community contacts. We identified eleven participants in the second group through snowball sampling. In each study site, we aimed to interview 15 community members: 10 participants in the first phase, and 5 in the follow-up phase. We aimed to achieve a balanced sample of age and gender among household participants. We considered this sample size to be a manageable workload for volunteers working during an epidemic and the Islamic holy month of Ramadan in the first phase, and an epidemic and rainy season in the second phase. After a preliminary analysis of data for each site, we revisited the need to increase the sample size to reach data saturation.

### Data collection

Semi-structured, in-depth telephone interviews were done by trained youth volunteers at a time convenient for consenting participants between May 10 and June 20, 2020, during the first round of data collection, and July 01 to August 10, 2020, in the second round. After obtaining participants’ permission, all interviews were audio-recorded. The interviews lasted between 40 and 70 minutes.

Interviewers used a pre-tested interview guide that contained open-ended questions. The interview guide covered the following topics: (1) knowledge of high-risk groups and protective measures for them, (2) the acceptability and feasibility of each of three shielding scenarios: (i) all extra-household activities are delegated to one family member who lives in a separate part of the house from the rest of the family, (ii) the high-risk person voluntarily shields, alone or with a carer, in a separate part of the house, (iii) neighbouring households or members of an extended family voluntarily ‘house-swap’ and group their high-risk members into dedicated houses, and, finally, (3) preferred and trusted sources and communication channels for information on COVID-19. Finally, participants were asked to suggest alternative or improved shielding strategies that would be suitable for their families or communities, and effective measures for promoting and implementing shielding in their communities. Interviews were conducted in Arabic.

### Data management and analysis

Audio-recorded interviews were transcribed verbatim by youth volunteers. We analysed data using a content analysis approach in two stages. The first stage was a participatory analysis through focus group discussions with the interviewers to generate preliminary ideas for transcript analysis, gain a thorough understanding of the context of data collection and identify immediate insights to assist responders. After the interviews were completed, interviewers were asked to listen to their recordings, familiarise themselves with the data and complete a summary report, summarising key ideas that emerged in conversations with participants. The interviewers were then invited to participate in group discussions with co-authors (SAEA, NA, NN, MD, IZ, AA), who used the reports to prompt follow-up questions and make use of the interpersonal dynamic of group discussions to generate new insights. These groups discussions were recorded and used by the co-authors to develop a y report of preliminary findings for quick dissemination to Y-PEER Sudan, the Ministry of Health and other COVID-19 responders.

The second stage was a thematic analysis using the transcripts. Data for each site was independently analysed for possible themes by the co-authors (NA, SAEA, NN, MD, IZ and AA). Themes were derived from the interview guide (deductive approach), by allowing emergent themes from the data (inductive approach) [20]. The analysis was then collated by the second author (SAEA) in a single spreadsheet. Through discussion amongst co-authors, consensus on themes and subthemes was reached, and categorisation was agreed. The themes were interpreted further and compared to the objectives of the study to generate the conclusions. Illustrative quotations were extracted by SAEA, translated into English by NA, and back-translated to Arabic by MAF. This article adheres to the Consolidated Criteria for Reporting Qualitative Research (COREQ) reporting guidelines [21].

## Results

We interviewed 89 participants: 59 during periods of movement restrictions and 30 participants after movement restrictions were eased (Table 1). The first group consisted of 25 females and 24 males, with an average age of 34 years old (age ranging from 20 to 70 years). The second group of participants consisted of 25 males and 5 females, with an average age of 44 years old (age ranging from 20 years to 83 years).

**Table 1.**
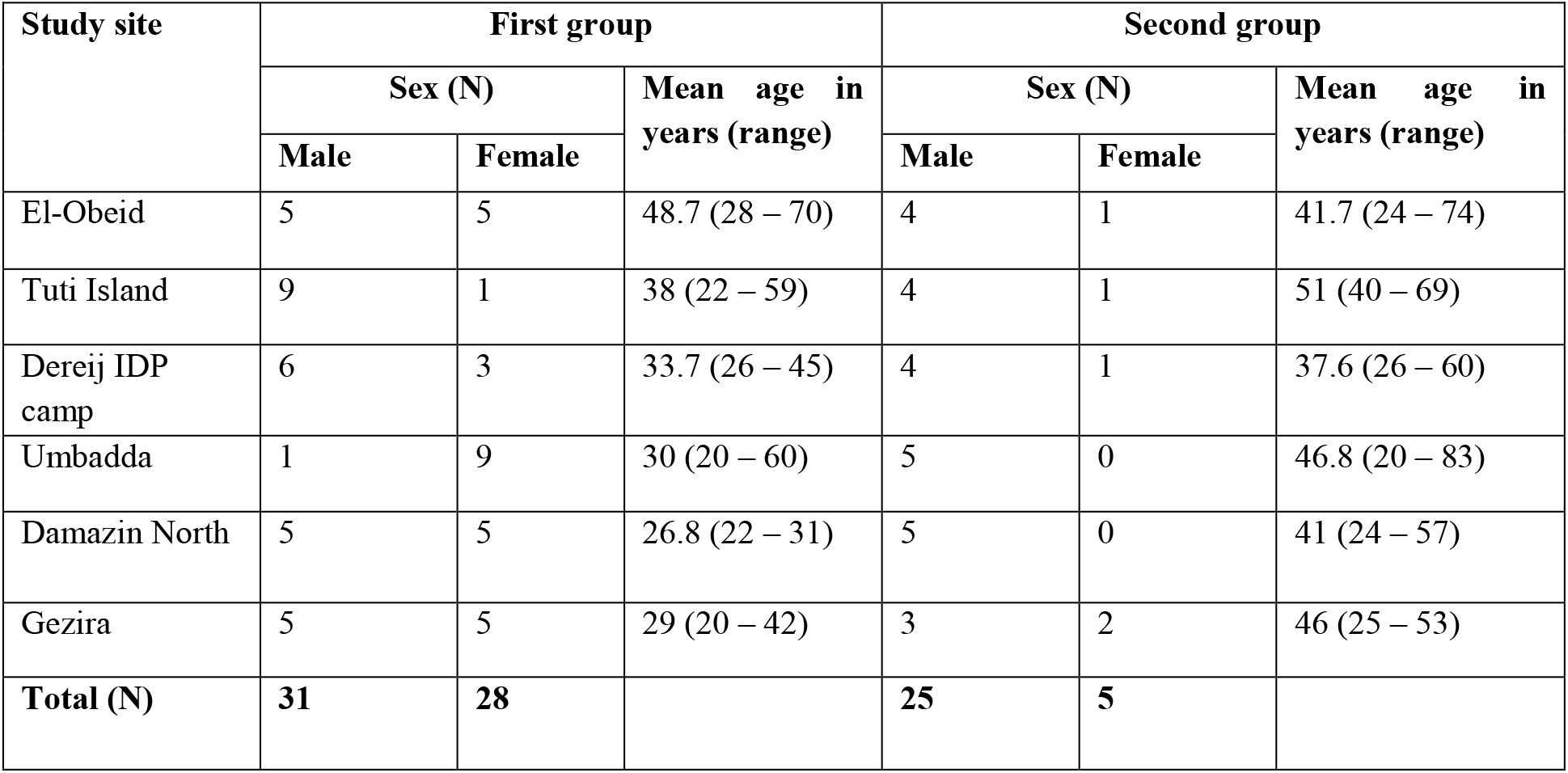
Age and sex of study participants (N=89)

| Study site | First group |  |  | Second group |  |  |
| --- | --- | --- | --- | --- | --- | --- |
|  | Sex (N) |  | Mean age in years (range) | Sex (N) |  | Mean age in years (range) |
|  | Male | Female |  | Male | Female |  |
| El-Obeid | 5 | 5 | 48.7 (28 – 70) | 4 | 1 | 41.7 (24 – 74) |
| Tuti Island | 9 | 1 | 38 (22 – 59) | 4 | 1 | 51 (40 – 69) |
| Dereij IDP camp | 6 | 3 | 33.7 (26 – 45) | 4 | 1 | 37.6 (26 – 60) |
| Umbadda | 1 | 9 | 30 (20 – 60) | 5 | 0 | 46.8 (20 – 83) |
| Damazin North | 5 | 5 | 26.8 (22 – 31) | 5 | 0 | 41 (24 – 57) |
| Gezira | 5 | 5 | 29 (20 – 42) | 3 | 2 | 46 (25 – 53) |
| **Total (N)** | **31** | **28** |  | **25** | **5** |  |

We present the findings by the following themes: knowledge about groups at high risk of severe illness or death due to COVID-19, knowledge and implementation of protective measures for high-risk groups, knowledge and implementation of strategies to shield high-risk persons, acceptability and feasibility of shielding strategies during periods of severe movement restrictions, participants’ recommendations to make shielding more feasible for communities, acceptability and feasibility of shielding strategies after severe movement restrictions were eased, and participants’ opinion on effective methods for promotion of shielding in their communities.

### Knowledge about groups at high risk of severe illness or death due to COVID-19

Most participants were aware that older people and those who have chronic diseases are at high risk of severe illness or death from COVID-19. We observed that knowledge levels were high across study settings, i.e. urban, rural and displaced populations.

*“I expect that if an elderly person with a chronic disease like hypertension or asthma, God forbid, gets COVID-19, it will be very difficult to prevent them from dying” –* male, 43 years, Dereij IDP camp

One participant observed that all COVID-19 deaths in her community were amongst older people with chronic diseases.

*“To be honest, from my observations, those who died [from COVID-19] all had chronic illnesses. They were elderly and had chronic diseases. For example, they would get a normal fever and vomiting, and in less than a week, maybe three or four days, they die. I now know that people with chronic illnesses don’t experience long COVID-19 illness before they die” -* female, 35 years, Dereij IDP camp

Participants reported multiple sources of this information, including radio, television, social media pages and groups, mass text messages, leaflets and other community members.

### Knowledge and implementation of protective measures for groups at high risk of severe illness or death due to COVID-19

To protect people at high risk, a few participants mentioned that those living with them should wash their hands and change their clothes when returning home to avoid transmitting the infection within the household. However, the majority of participants reported measures for those at high risk themselves, such as maintaining their general health and well-being and reducing social contacts outside the household.

*“I have a special diet for [my father], and I give him vitamin C from time to time to boost his immunity, these are the things that I managed to do. I manage to keep him at home because he is elderly. For the rest of the family members, I look after them too, but I give him special care because he is the oldest person at home and is elderly, so he can be vulnerable to anything”* – male, 28 years, ElObeid

To reduce social contacts, participants mentioned advising older people to reduce or abandon social visits and gatherings, wear face masks and abstain from physical greetings outside the home. Other participants mentioned supportive measures to help those at high risk stay at home. For example, in El-Obeid, a participant reported that there was a local community-based initiative called “Dawak Fi Beitak” which arranges regular home delivery of medications to those with chronic illnesses.

Nonetheless, participants reported challenges to compliance with these protective measures. These include low risk perception of COVID-19, COVID-19 denial, sociocultural barriers and financial challenges.

*“The most important thing is that a person [who is at high risk] is convinced that COVID-19 is dangerous to them. They must be aware of the danger of COVID-19 so they can protect themselves and others”*. - male, 54 years, Tuti Island

Another participant explained how social desirability and cultural traditions make it difficult to comply, particularly by the elderly, who are revered in Sudanese culture.

*“Our culture can [become an obstacle]. Even though the seriousness of this disease is communicated through all media, people say that they cannot refrain from physical greetings, as these are cultural habits and traditions. This kind of attitude makes you worry [about infection with COVID-19]. No matter how often we advise elderly people to be isolated at home, it is difficult to implement this in our community*.*” -* male, 31 years, Dereij IDP camp

Other participants reported that high-risk family members insist on maintaining their social relations by welcoming visitors or visiting others, continuing to pray at the mosque and attending important social events such as funerals.

*“In our community, people go to Friday prayers since mosques are still open even though they are supposed to be closed. So, people continue to go to Friday prayers and evening prayers in Ramadan. I tried to stop [my mother] from going, but she also refused because when she goes to pray in the mosque, the people there tell her that prayers will end the epidemic and when we pray in the mosque [COVID-19] will not infect us, can you believe that? So, they [should close these mosques]*.*” -* female, 31 years, Umbadda

In the rural study site, participants explained that most young family members are away working in cities, leaving the elderly with the responsibility of running households.

*“You will find that many households don’t have young people, especially in rural areas, as they go to the capital city or other areas [where there are jobs]. You rarely find a home with 2 or 3 young people; they are usually elderly above 50 years or children under 15 years - so the elderly person has to leave home to provide the household’s needs*.*”* - male, 28 years, Gezira

Unlike other study sites, none of the participants from Damazin North reported implementing measures to protect family members at high risk.

### Knowledge and implementation of strategies to shield those at high risk of severe illness or death due to COVID-19

Although most participants had good knowledge of high-risk groups and mentioned several protective measures, the shielding concept was new to them. Many participants confused it with isolation or quarantine and interpreted it as a complete disconnection from the family and community. There was also confusion as to whether shielding occurred for those that have COVID-19-like symptoms or as a prevention measure. Nonetheless, four participants were already implementing some form of shielding at home at the time of the study.

*“I am currently using [shielding] at home with my father; I have set him up to stay in a separate part of the house, a separate room. I provide him with everything he needs, and he prays in there too, he doesn’t leave the house*.*” -* male, 28 years, El-Obeid

### Acceptability and feasibility of shielding strategies during periods of severe movement restrictions

For this study, we suggested and discussed three shielding strategies with the respondents to explore their opinions on their acceptability and feasibility. The strategies are: (i) all extra-household activities are delegated to one family member who lives in a separate part of the house from the rest of the family, (ii) the high-risk person voluntary shields, alone or with a carer, in a separate part of the house, (iii) neighbouring households or members of an extended family voluntarily ‘house-swap’ and group their high-risk members into dedicated houses.

### Delegation of extra-household activities to one family member who lives in a separate part of the house from the rest of the family

In general, this strategy was acceptable to many participants, either fully or partially.

*“This is a very good suggestion. I am now using this strategy and applying it at home. I am the only one in the family who leaves the household for shopping – I bring anything that is needed. No one else leaves the house*.*” -* male, 28 years, El-Obeid

Although many agreed that it is feasible to choose one household member for all extra-household activities, most were not convinced that isolating that person within the household is possible or necessary. Instead, most participants emphasised that the delegated person should adopt infection prevention measures when returning home, such as washing hands, changing clothes or bathing.

*“This first suggestion is the best one. Each household selects one family member to provide all their needs, but that person should not isolate themselves – when they come from outside, they can put their clothes in the sun, and wash their hands with water and soap”* – male, 22 years, Tuti Island

*“My husband always goes out to bring our household needs. He is very careful not to harm anyone or a child at home if he is feeling unwell or sick, but we have never isolated him, and we never will – we will not leave him alone” -* female, 43 years, Umbadda

One participant reported concerns about isolation resulting in the stigmatisation of the delegated person:

*“This is a good suggestion but difficult to implement. We live in a community where social interaction is intense, and so isolation is difficult. The feeling of isolation may make the person feel stigmatised and trigger thoughts that they may actually be infected. This [suggestion] requires a great degree of awareness, insight and acceptance – it can be implemented but not across all of Sudan*.*” –* male, 31 years, Damazin North

Another participant noted the difficulty in adhering to isolation within the household, mainly because the delegated person is healthy.

*“We as Sudanese are very close and intimate [within our families]. If I ask you, with your knowledge and work in this field of COVID-19, to stay away from your father or other household members while you are sick, you can stay away from them because of their safety so they [do not get infected]. But if you are well, you will not be able to do it. Inadvertently, you will come close [to your family members]*.*” –* Male, 28 years, Damazin North

Participants who did not accept this strategy mentioned several reasons, particularly the difficulty in having the household rely on one person’s income.

*“To have one person from the household leave the house – you know that one person cannot [financially] support a household. From each household, approximately three people are working to cover the household’s expenses – it is difficult to rely on one person*.*”* - male, 46 years, El-Obeid

Also, people within households play different roles and have different responsibilities, particularly in households of extended families, and in rural areas.

*“It’s difficult. At home, some people are working on farms, others in the market. Also, most of these people have limited income, and a person with limited income cannot stay at home*.*” -* male, 26 years, Gezira

### Voluntary shielding of the high-risk person in a separate place within the household

For many participants, this strategy was acceptable primarily because it is in keeping with the setting of the Sudanese home.

*“I agree with this strategy, absolutely. And this already exists – in families with an older person, such as a grandfather or father, they have their room or their hut, and other members of the household serve them with food, or help then bathe. I agree with this suggestion totally*.*” –* male, 43 years, Dereij IDP camp

Participants mentioned several implementation challenges. For most, there are social challenges to isolating an older person from the rest of the household, related to the respect and reverence of parents and the elderly. Some reported that they cannot imagine their household life without the involvement of their parent or elderly family member, especially for family activities such as meals, and that they were concerned about the impact of isolation on the shielded person. Others reported that they would be concerned about what the rest of the community would say about them if they isolated their elderly family member.

*“My children will never accept their mother isolating in a separate room. We are all taking precautions and being careful, we are relying on Allah [to protect us]” -* female, 43 years Umbadda

*“This isolation that you mention, I think it’s difficult. I, for one, cannot eat alone. People are used to eating together, spending time with each other and chatting together [as a family]” – male, 54 years, Tuti Island*

Other participants reported that the high-risk family member would refuse to be isolated, even if the rest of the family accepts it. This refusal is somewhat linked to the lack of awareness of personal risk by the elderly themselves – several participants reported that older people were not as aware as others of the risks of COVID-19.

A few participants, particularly in Dereij IDP camp and Umbadda, reported a lack of space within low-income households to facilitate shielding.

*“If I was able to implement this strategy in my home, other people will not be able to, due to limited spaces in the camp. I can implement this in my family and home, but my neighbours can’t” –* female, 35 years, Dereij IDP camp

As with the first strategy, participants suggested the use of prevention and safety measures when dealing with elderly or high-risk persons at home, such as masks and handwashing, as an alternative to isolation.

### Shielding of high-risk persons outside the household in a voluntary ‘house swap’ arrangement

In this strategy, where there is not enough space for shielding within the home, neighbours or extended families voluntarily swap houses to give high-risk individuals and their carers a separate house to shield in, and a house for those at low risk to live in and continue to work. For most participants, this strategy was neither acceptable nor feasible.

*“No one will accept to leave their house no matter what. Even if there is [an epidemic], they will tell you that they can only be comfortable in their own home” -* female, 27 years, Gezira

Respondents explained that households have different habits and rituals within their homes and that elderly family members need the protection of their family and the comfort of their own homes for their well-being.

*“This cannot be implemented because traditions are different; even people’s habits are different. For example, the type of food that is presented to a [high-risk] person at home may be different from other family members. You can manage to do this within a household, but not in an extended family, like relatives or neighbours. If a [high-risk] person needs special food, but they are [living] in one place with other [high-risk] people, they will all be served the same food, so there will be compromises in this area some elderly people need someone to bathe them or feed them so if I put them with other people that have not dealt with them before, there will be compromises [in their care*]” -male, 28 years, El-Obeid

Older participants confirmed this unwillingness to leave their homes, citing a lack of trust in others, the importance of maintaining traditions, and fear of criticism from the surrounding community.

*“[Shielding outside the home] cannot be done. You know, Sudanese people have changed, they are not [trustworthy] like they were before, and the economic situation has also [worsened]. For me, I don’t think it will happen. I cannot leave my home and stay in another person’s house. As you know, Sudanese people talk – people in Sudan cannot understand what is happening [with the epidemic]” -* female, 63 years, El-Obeid

A few participants agreed that this strategy might be acceptable as it builds on the communal solidarity prevalent in Sudanese communities. This exception was most pronounced in Dereij IDP camp, where half of the respondents thought the strategy was acceptable. Nonetheless, they reported that this would require a high level of awareness of risk in the population, and is more likely to be acceptable in a house swap with close relatives.

*“This is acceptable, very acceptable. We as Darfuris are renowned for our generosity; people are very cooperative with each other -* male, 26 years, Dereij IDP camp

### Participants’ recommendations to make shielding more feasible for communities

Several respondents suggested measures to help those at high-risk to stay at home, including the delivery of regular medications and home-based clinical monitoring services. Others emphasised the importance of maintaining access to health services for the elderly and those with chronic illnesses during the COVID-19 epidemic.

To support at-home shielding, many participants suggested the provision of financial and in-kind support, especially for poor households, to enable families with high-risk members to shield at home and comply with prevention measures. For instance, participants suggested the provision of food baskets, soap, gloves and hand sanitiser. Expectations of external assistance were generally higher in Dereij IDP camp compared to other study sites.

*“Older people and people with [chronic] diseases should be more careful when dealing with other people; they should always be using hand sanitisers. This is the only problem I see. [Neighbourhood committees] can provide these people with [material] assistance [at home] so they don’t have to go to work, as mixing in the market [with others] can transmit the disease. Some people are convinced that COVID-19 exists, and other people say they would rather die of COVID-19 than starvation*.*”* –female, 23 years, Umbadda

Participants also mentioned that COVID-19 transmission in the general population should be reduced, and suggested increasing information dissemination through the media, provision of advice from healthcare workers to local communities, and promoting preventive behaviours in public areas. Participants in rural areas also requested better access to COVID-19 testing as testing centres were only available in urban areas. Some participants also suggested better enforcement of policies, such as lockdowns and inter-state travel restrictions, to create an environment that is more supportive of shielding.

*“I think that the policy has to be comprehensive. For example, during the lockdown in Khartoum or El-Obeid or other places, people are not complying, and the authorities are not enforcing the implementation, commercial activities should be suspended now. My father is one of the people who is not complying with the lockdown; he goes to work. If there was serious enforcement of the lockdown, my father would have complied and stayed at home, and I would be going out to get the household’s essential only. I would be implementing this [shielding] strategy you mentioned. But I cannot implement it now if there aren’t other measures implemented, it has to be a complementary policy - I cannot implement one part and ignore the other”* – Male, 32 years, El-Obeid

### Acceptability and feasibility of shielding strategies after severe movement restrictions were eased

In general, the acceptability and feasibility of shielding strategies were reduced after movement restrictions were eased, as participants reported lower perception of risk in their communities. Some participants mentioned that, in addition to widespread beliefs that COVID-19 transmission in communities no longer exists, several rumours and misinformation about COVID-19 were prevalent in their communities, and COVID-19 is no longer at the forefront of their concerns or interest. In the absence of risk awareness, it was unlikely that community members would consider taking up shielding.

*“In the current moment in our community, theoretically shielding is possible and if people are convinced to take it up, it is possible. But there are many issues we didn’t discuss that have to do with awareness. Fear of the virus in the community I live in has regressed, and this means that even effective strategies [will not] be adopted, because people will only adopt it when they feel at risk, and the risk has regressed a lot. Also, there are rumours and misbeliefs in the community, about whether the virus exists, people are not giving it much attention. So, the idea of isolating the elderly or an ill person is possible but what can prevent it is people’s awareness – there are very few people who are aware” -* male, 24 years, El-Obeid

*“In [our] neighbourhood community, it is not acceptable that a person does not leave their house [nowadays]. Some people are saying there is no COVID-19. [Shielding] may be acceptable in communities that are aware, for example, in Khartoum neighbourhoods that have high awareness or other [similar] communities. There, you can implement this, and it is an effective [strategy]”* - male, 83 years, Umbadda

Challenges to implementation of shielding were similar to those mentioned by participants interviewed during the period of movement restrictions, including socially-driven unwillingness to segregate family members, particularly the elderly, and financial losses as a result of shielding. However, we noticed that compliance with social commitments outside the household, such as visits to family, funerals and weddings, became more pressing after movement restrictions were eased.

*“All these [shielding strategies] are good. But people cannot implement them in our community. Community members are now mingling and want to socialise with each other. They cannot leave our traditions. Our traditions are the problem*.*” -* male, 54 years, Gezira

### Participants’ opinions on effective communication and promotion of shielding strategies in study communities

Many participants mentioned that diverse mass communication channels should be used to broadcast information widely, including television, radio, social media, text messages and displaying visual materials in public places. Different channels reach different population groups. For example, social media is widely used by younger community members, especially in urban areas, whilst radio still plays a significant role in rural areas where internet coverage is low and televisions not widely present. Within their communities, many participants mentioned the use of mobile activities, such as vehicles with megaphones, and edutainment activities such as theatre.

*“The source we use most frequently is Facebook, and also radio, but mostly Facebook and WhatsApp, especially among young people. Older people and rural areas [use] radio and television. They also listen to the local ‘sheikh’ [religious leader] because they go to him with their problems so that he can raise their awareness [of COVID-19]” –* female, 20 years, Umbadda

To complement mass communication, many participants mentioned the importance of using local, trusted persons to promote shielding. Influential community figures mentioned by participants included members of neighbourhood committees (primarily youth), religious leaders, teachers, local health workers and other community leaders, such as members of the local civil administration. In urban settings, local health workers, neighbourhood committees, religious leaders and family members were the most trusted persons, whereas in rural areas, family members, especially those working in the medical field, were influential. In the IDP camp, local community and religious leaders were the most credible sources of information. Some participants mentioned that trusted persons could use their existing platforms to share information, for example, imams giving sermons in mosques. Other participants suggested more in-person activities such as household visits with infection prevention precautions to families with high-risk family members.

Several participants mentioned the importance of using local dialects and languages to reach all groups within the community, particularly the elderly, as many mentioned that current mass communication activities use formal language that is not widely understood or preferred by their communities. Other participants commented that some activities are inappropriately designed; for example, one participant mentioned that mobile megaphones travel too fast to be fully heard or operate at inappropriate times of the day.

*“I think awareness activities need to be continuous, especially in rural areas who may not have a television or radio. Awareness-raising activities using simple means can be implemented there, and they can use local dialects. Many [elderly] people now hear the megaphone from the street, and they ask me “what are they saying?” and I have to explain to them what is being said about COVID-19” -* female, 32 years, Dereij IDP camp

Some participants recommended that positive messaging and language should be used to promote preventive behaviour, and to avoid promoting preventive behaviour by instigating fear in the population, through language such as “the virus will kill you…”. Other participants recommended that high-risk community members need to be targeted with tailored risk communication to increase their understanding of their risk of severe illness and death, and to empower them to take measures, such as shielding, to protect themselves.

## Discussion

In low-resource settings, prevention is key to mitigating the impact of COVID-19. Shielding of high-risk individuals, either within their homes or in safe communal structures, has been suggested as a realistic alternative to widely-enforced social distancing measures such as lockdowns that may cause severe economic repercussions [22]. While shielding is premised on sound epidemiological principles and has shown through modelling to cause significant reductions in hospitalisations and mortality due to COVID-19, wide-scale implementation of shielding can be fraught with social, cultural and operational challenges. In this study, we explored the acceptability and feasibility of shielding strategies during the COVID-19 epidemic in six illustrative communities in Sudan. We elicited contextual adaptations for the implementation and promotion of shielding in the study communities.

Across the different study settings, the majority of participants knew that some individuals were at higher risk of severe COVID-19 outcomes, particularly the elderly. This high level of knowledge may be indicative of effective dissemination of information about COVID-19 through multiple channels across the study sites. Similar levels of adequate knowledge of groups at higher risk of severe COVID-19 outcomes were reported in other low- and middle-income settings, including Kenya and Egypt [23, 24]. Despite that, only a few participants knew of measures to reduce intra-household contacts with persons at high risk. Many participants initially confused shielding with quarantine or isolation, and further information was needed to clarify the difference between these measures – similar misunderstandings were reported by a study in Goma in the Democratic Republic of the Congo [25]. Study participants were, however, positive when presented with concrete options for protecting high-risk family members within the household.

Shielding of high-risk individuals was generally acceptable, and many participants’ remarks referred to strong Sudanese traditions of protecting the elderly and the vulnerable. Consultations with crisis-affected communities in Yemen also reported acceptance of shielding within the context of strong cultural tendencies to protect the elderly and most vulnerable [25]. Similarly, a shielding intervention in Ethiopia leveraged a culture that values community solidarity and cohesion, where communities traditionally prioritise those most in need of protection [26].

Our findings indicate that there was no outright acceptance or rejection of any of the proposed shielding strategies. However, study participants were on a broad spectrum in terms of readiness to implement shielding. In general, shielding strategies within the household were acceptable, while shielding outside the household was widely unacceptable due to concerns about privacy, cultural norms and compromises in caregiving for shielded family members. Consultations by humanitarian actors with displaced communities in Yemen and Lebanon yielded similar findings of strong preference of household-based shielding and an aversion to shielding outside the household [27]. In Yemen, community members found shielding within the extended family acceptable where extended families live in compounds of neighbouring houses [25]. In our study, participants from the displacement camp were more accepting of extra-household shielding; interviews with displaced Syrians in opposition-controlled camps in Northwest Syria found a similar willingness to consider house swapping arrangements for shielding [28].

We also found that acceptability of shielding is not static, but is influenced by contextual changes, where enthusiasm for shielding had diminished in study communities after severe movement restrictions were eased. The new context was one where a change in government strategy fuelled widely-prevalent beliefs that the epidemic had indeed passed and created increased pressure to comply with social commitments. These findings are corroborated by previous research, which showed that both government response measures and perceived risk of the severity of respiratory infection epidemics had a significant influence on behavioural intentions towards social distancing [29-31].

In all our study communities, feasibility was a determinant of acceptability. Feasibility was challenged both socially and financially by study participants. Among the elderly, the loss of social capital due to eliminating social contacts outside the home, especially in mosques and social gatherings, was a significant barrier to the feasibility of shielding. Within the household, participants had concerns about the implications of segregation within the household on family well-being, with some suggesting less strict separation combined with infection and prevention measures such as maintaining hand hygiene within the household. Furthermore, a few participants raised doubts about the sustainability of full compliance with segregation within the household, particularly where none of the family members was unwell. These findings indicate that while shielding is widely acceptable, the effectiveness of shielding in these communities may be lower than predicted by mathematical models, which assume drastic reductions in contacts within the household [15]. Furthermore, this may indicate an intention-behaviour gap amongst participants who accept household-level shielding but question their ability to implement or sustain it. Previous researchers have also shown that low self-efficacy attenuates behavioural intentions and behaviour adoption in the context of epidemics of respiratory infectious disease [29, 31, 32].

Our findings also show that a shielding promotion campaign requires a nuanced approach that addresses different COVID-19 beliefs and perceptions, including those who do not believe COVID-19 exists in their community or at all. Communication channels should combine mass communication with personal communication from trusted and influential figures in the local communities. Our study participants report that the elderly tend to be least informed about COVID-19 and their levels of risk, and are less likely to comply with preventive behaviours in general. Other studies in Ethiopia and Bangladesh also found lower levels of knowledge about COVID-19, and inadequate compliance with preventive behaviours among the elderly and individuals with chronic illnesses [33-35]. These findings indicate that shielding promotion requires a targeted risk communication strategy for those at high risk of severe COVID-19.

Our study has limitations. Firstly, in this study, we used interviews to explore the acceptability and feasibility of shielding. In acceptability research, focus group discussions are preferred over interviews as they have the advantage of allowing group members to collectively brainstorm and debate ideas, opinions, and recommendations [36]. However, given the movement restrictions posed by COVID-19, remote interviews were the appropriate choice for our study, and we are confident that we reached data saturation, as no additional themes emerged in the final interviews at any of the study sites. Secondly, we conducted the interviews remotely by telephone, which is subject to challenges including challenges to establishing rapport, absence of visual cues and contextual data, and the relatively short time of the interview to avoid respondent fatigue [37-39]. Thirdly, in this study, we inquired about the acceptability of a new concept of which the majority of study participants had no prior knowledge. It is possible that some participants provided their opinion without considering the implications of shielding on all aspects of their lives. However, given that reasons for and against acceptance were recurrent across participants and study sites, we are confident that this limitation had a minimal effect on the findings, probably in the direction of acceptance. Finally, given our study design, findings cannot be generalised to all Sudanese communities.

Shielding promotion in the study communities should provide realistic options and practical guidance for minimising social contacts within households, and should be accompanied by financial and social interventions to support feasibility. Financial assistance by the Government of Sudan and other donors should be directed to interventions that support shielding, such as providing home-based treatment for shielded individuals, and direct financial support for families, including where the breadwinner is at high risk of severe COVID-19 outcomes. Interventions targeting shielding families and the larger community are needed to address concerns about psychological well-being and social isolation of shielded individuals. In the former category, shielding promotion should adopt an approach that highlights and leverages local traditions for protecting and the elderly and most vulnerable in times of hardship, and interventions for incentivising and celebrating local community action that supports shielding. For individuals and families that are shielding, supportive interventions may include psychological health guidance and support, distribution of shielding aids such as soap, face coverings and cleaning supplies, and innovative remote socialisation methods. To provide a supportive environment for shielding, shielding promotion should occur in the context of broader communication interventions and policy responses that enhance risk perception of COVID-19, address misinformation and incentivise compliance with preventive behaviours in the population. Finally, to improve uptake, shielding promotion should be time-limited, by actively promoting the strategy when there are early indicators of increasing incidence of COVID-19, and reducing promotion when incidence and risk of COVID-19 are deemed low by responders.

## Conclusion

Our study findings provide valuable insights and recommendations for COVID-19 responders in Sudan and similar resource-poor settings. We conclude that shielding is generally acceptable in the study communities, and that acceptability is influenced by feasibility, and by contextual changes in the epidemic and associated policy response. Shielding promotion interventions in Sudanese communities may capitalise on the cultural sense of duty towards elders and vulnerable groups and should address different COVID-19 beliefs and perceptions. Promotion campaigns should include targeted risk communication and shielding promotion amongst individuals at higher risk of severe COVID-19 outcomes. Households should be provided with practical guidance to implement realistic household-level shielding options, with social, psychological and financial support interventions to sustain shielding effectively. Furthermore, communities should be incentivised and rewarded for supporting and enabling shielding.

## Supporting information

Reporting Checklist (COREQ)

## Data Availability

The datasets generated and analysed during the current study are available from the corresponding author on reasonable request.

## List of abbreviations

COREQ: Consolidated Criteria for Reporting Qualitative Research
COVID-19: Coronavirus disease
WHO: World Health Organization
Y-PEER Sudan: Sudan’s Youth Peer Education Network

## Declarations

### Ethics approval and consent to participate

The ethical approval for the study was obtained from Sudan’s National Health Research Ethics Committee based at the Federal Ministry of Health (number 1-4-20) and the London School of Hygiene and Tropical Medicine’s Research Ethics Committee (number 22051). All participants provided verbal informed consent, including explicit permission to record the interview and to publish anonymised quotes from the interview. Participation was voluntary, and participants were not financially rewarded for participation.

### Consent for publication

Not applicable

### Competing interests

The authors declare that they have no competing interests

### Funding

The data collection costs were borne by the investigators and volunteer youth peers. NA was supported by UK Research and Innovation as part of the Global Challenges Research Fund, grant number ES/P010873/1. Article Processing Charges were covered by UK Aid via The British Council, as part of the ‘Community-led COVID-19 Mitigation in Sudan: A Research and Advocacy Intervention’ project. None of the above funding bodies had any role in the design of the study and collection, analysis, and interpretation of data and in writing the manuscript.

### Authors’ contributions

NA, NN and MD conceptualised the study and developed data collection tools in discussion with other co-authors. IZ and AA coordinated and supervised data collection. SAEA, NA, NN, MD, IZ and AA analysed and interpreted the data. NA and SAEA drafted the manuscript and NN, MD, IZ, AA, and MAF read, reviewed and approved the final manuscript.

## Acknowledgements

We are grateful to the youth peers for their critical role in data collection and preliminary analysis, and for their valuable insights into the context at the study sites. We thank Dr Jennifer Palmer and Professor Bayard Roberts for the valuable comments and ideas on the study conceptualisation and design. We thank Ms. Sian White for her assistance with drawing out behavioural insights from study findings, and shaping recommendations for shielding promotion in Sudanese communities.

## Notes

### Competing Interest Statement

The authors have declared no competing interest.

